# Modeling the Spread and Control of COVID-19

**DOI:** 10.1101/2020.09.16.20195826

**Authors:** Ashutosh Trivedi, Nanda Kishore Sreenivas, Shrisha Rao

## Abstract

Data-centric models of COVID-19 have been tried, but have certain limitations. In this work, we propose an agent-based model of the epidemic in a confined space of agents representing humans. An extension to the SEIR model allows us to consider the difference between the appearance (black-box view) of the spread of disease, and the real situation (glass-box view). Our model allows for simulations of lockdowns, social distancing, personal hygiene, quarantine, and hospitalization, with further considerations of different parameters such as the extent to which hygiene and social distancing are observed in a population. Our results give qualitative indications of the effects of various policies and parameters; for instance, that lockdowns by themselves are extremely unlikely to bring an end to an epidemic and may indeed make things worse, that social distancing matters more than personal hygiene, and that the growth of infection comes down significantly for moderately high levels of social distancing and hygiene, even in the absence of herd immunity.

## 1 INTRODUCTION

The COVID-19 pandemic has presented humanity with grave social and economic challenges, and also a whole host of research questions that are not easily answered using standard statistical and other methods, in large part on account of the lack of sufficient data about the novel coronavirus SARS-CoV-2. Of particular interest is to understand emergent situations that may occur as a consequence of policy choices (e.g., lockdowns) and individual behaviors (e.g., observance of social distancing and personal hygiene). Policy-makers and the public at large can benefit by a better understanding of the macro-level consequences of the delicate micro-level behaviors of individuals. These macro-level consequences are emergent properties that arise from the interactions of a large number of diverse individuals who are part of the system.

Agent-based models (ABMs) help us consider such possible realities by creating and studying the effects of suitably designed agents in an interactive environment. Each agent represents an individual person with certain specific properties and behaviors. These agent properties are carefully chosen to model humans realistically, and agent behaviors are programmed keeping real behaviors in mind. With ABMs, it becomes feasible to study the complex system-level properties which arise from the interactions of individuals.

Certain specific aspects of our ABM for COVID-19 are a larger set of possible states that enable distinctions between asymptomatic and symptomatic infections, and parameters to model different levels of hygiene and social distancing. The model also allows for different levels of hospital care for differences in severity of illness. Unlike statistical models which often assume a homogeneous population, our model allows for agents with different levels of health, immunity, and comorbidity. Using simulations based on this ABM, we are able to evaluate different possible circumstances and observe their consequences.

Modeling of infectious disease outbreaks is an endeavor of long standing [24, 49], but almost all efforts use mathematical (equation-based) modeling techniques [7]. There have also been specific efforts at modeling possible flu-like pandemics caused by H5N1 (“bird flu”) [3] and H1N1 (“swine flu”) [26].

In case of the very real COVID-19 pandemic also, there have been numerous analytical efforts based on mathematical models [1, 4], as well as data-driven models and projections [16, 34].

Agent-based models have also been used in epidemiology [21, 33, 35, 47], and to model specific instances such as mosquito-borne diseases [22] and pandemic influenza [25]. Agent-based models have some advantages over mathematical models [20], such as in allowing for consideration of heterogeneity in the population, discovery of emergent properties that are not readily apparent from the system description, and study of behaviors for which no closed-form mathematical description is easily available.

There are also some early efforts at modeling COVID-19 using agent-based models [10, 44].

The present work is distinct in that we create an extended agent-based model that allows for qualitative evaluations of the effects of social distancing, personal hygiene, and lockdowns, while taking into account epidemiological characteristics of COVID-19 [12] and the heterogeneity of populations of people who may become affected. The basic SEIR model and its common extensions are not entirely adequate to model the spread of the disease, as they do not take into account the classification of infectious states by types—asymptomatic, symptomatic (but not hospitalized), hospitalized, and needing intensive care—that is a concern in reality [2].

In our model, each agent represents a human in society who interacts with others. Human movements and interactions are captured by motion and occasional proximity in a two-dimensional grid. Each agent has certain characteristics (see Section 3.2.1 for details), and some agents change states based on infections and recovery. Our model allows us to understand the difference between a black-box view of the epidemic and a glass-box view (Section 3.1). It also allows us to control agent parameters to accord with the characteristics of different people in a large human population (Section 3.2), and to change the characteristics of the system to model the effects of different policies and public-health practices (Section 3.2.2).

Our results are in line with expectations; e.g., it is suggested by Schueller *et al*. [42] that lockdowns slow down the spread of COVID-19 and reduce the peak of infections. Silva *et al*. [44] propose an agent-based model that allows the simulation of social distancing and the use of masks (as a specific measure of personal hygiene), and study their effects in certain scenarios, with similar but incomplete results.

The following are some of the findings of our work:

1. Social distancing matters more than personal hygiene (Section 4.3).
2. Social distancing and hygiene together matter more than lockdowns, for controlling the spread of disease (Section 4.4). In fact, in their absence a lockdown can hurt more than help—without social distancing and hygiene, a post-lockdown peak of cases can be higher than without lockdown, with a similar effect being also seen with more stringent lockdowns as compared to more relaxed ones (Figure 7).
3. When the fraction of the population that is immune to the disease (either by vaccination or by previous exposure or recovery) is small, say 20%, the peak of infectious cases can be nearly as high, in the absence of social distancing and personal hygiene, as when there is no immunity at all in the population (Section 4.5). Even in case some 40% of the population is immune, there can be a large number of infections in the absence of social distancing and hygiene (Figure 12b).
4. Proper herd immunity in the classical sense is said to occur (in the total absence of social distancing and hygiene) only when some large fraction of the population becomes immune [38]. However, when a smaller fraction of 40% of the population is immune (which we describe by saying the fractional immunity of the population is 0.4), this can be achieved with some observance of social distancing and hygiene; the same effect is also possible with an even smaller fraction of 20% of the population being immune, with a rigorous observance of social distancing and hygiene (Section 4.5).
5. Surges in hospital capacity (including critical care capacity) help reduce the number of severely ill people dying due to lack of access to medical care, but do not directly affect the overall number of cases to a significant extent (Section 4.6).

## 2 DISEASE MODEL

A common way of modeling the spread of infectious diseases is using the SIR (susceptible-infectious-recovered) model [49], which is a *compartmental* model that divides the entire population into three classes. The *susceptible* state consists of individuals who may become infected; the *infectious* individuals are those currently afflicted with the disease; and *recovered* individuals are ones who have overcome the disease.

An extension of this is the SEIR model [49] that also allows for an *exposed* state where an individual has received a load of the pathogen, but is not yet infectious or symptomatic. In the SEIR model it is generally assumed that recovered individuals have lifelong immunity.

Both the SIR and SEIR models are generally based on differential equations, and allow for various types of qualitative analyses [14, 46].

We modify the usual SEIR model to also include multiple infectious states, and to also show when someone is deceased.

### 2.1 Modified SEIR

The primary motivation for this modification is the observation that in COVID-19 (but also in other diseases) the infectious state is not truly a single state describing all infected individuals. Rather, there are multiple types of infectious states:

*I*_0_ This is the *infectious asymptomatic* state, where an individual does not show signs of the disease at all, but can transmit it to others. It is presently believed that a large number of infected people recover directly from this state, and that coming into proximity with such asymptomatic individuals is responsible for a large fraction of the COVID-19 spread [6].

*I*_1_ This is the *infectious symptomatic* state where an individual shows signs of the disease, but is not (yet) hospitalized, either because the symptoms are considered mild enough, or because hospital care is unavailable.

*I*_2_ This is the *infectious hospitalized* state, where an individual shows signs of the disease and receives hospital care.

*I*_3_ This is the *infectious critical care* state, where the individual is severely afflicted and receives critical care in an ICU or similar, including with specialized devices such as ventilators.

In addition to this, we add a distinguished state *D* to the model, showing when a sick individual is deceased as a consequence of the illness. The modified SEIR model is shown in Figure 1.

**Figure 1:**
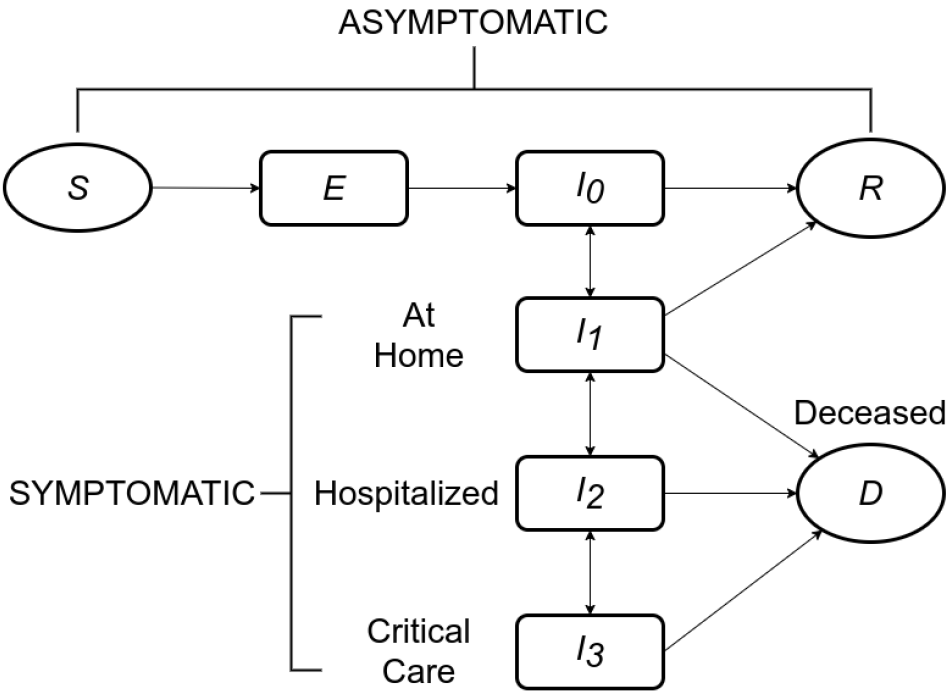
Modified SEIR model for COVID-19, with multiple infectious states.

This model is slightly similar to the one proposed by Arino *et al*. [4] which also considers multiple types of infectious states, but that model is based on differential equations and thus not directly applicable in an ABM context.

### 2.2 State Transitions

The state transitions between different states for an individual are also as indicated in Figure 1.These are as follows.

*S* → *E* The state transition from *susceptible* to *exposed* happens when a susceptible individual comes into close contact with an infectious individual (someone in any of the states *I*_0_, *I*_1_, *I*_2_, or *I*_3_). Each infectious individual has an infection range, and if another individual comes within their infection range, they are said to be in close contact. The state transition from *S* to *E* is considered instantaneous (see Section 3.3.1).

*E* → *I*_0_ The state transition from *exposed* to *infectious asymptomatic* happens upon the completion of the *latent period* of the disease, starting from the instant of exposure (see Section 3.3.2).

*I*_0_ → *I*_1_ The state transition from *infectious asymptomatic* to *symptomatic* happens upon the completion of the *incubation period* of the disease, starting from the instant of exposure. In many individuals, this transition may not happen at all, and the individual may instead transition to *R* (*recovered*) directly (see Section 3.3.2).

*I*_1_→ *I*_2_ The state transition from *infectious symptomatic* to *hospitalized* happens in some infected individuals when the symptoms worsen to the point of requiring hospital care (see Section 3.3.3).

*I*_2_ → *I*_3_ The state transition from *hospitalized* to *critical care* happens in some individuals whose aggravated conditions require ICU and similar critical care (see Section 3.3.4).

*I*_*_ → *D* The state transitions from *I*_1_, *I*_2_, and *I*_3_ to *D* (*deceased*) happen in some individuals. Individuals who are in *I*_1_ and *I*_2_ in need of more advanced care (in *I*_2_ and *I*_3_ respectively) are at increased risk of transition to *D* when such care is unavailable. Individuals in *I*_3_ are at increased risk of transitioning to *D* with increasing time spent in that state (see Section 3.3.5).

## 3 AGENT MODEL

In this section, we describe our agent model and its workings. In Section 3.1, we first encapsulate the difference between black-box and glass-box views of the spread of COVID-19, which is so important to understanding why this pandemic has proven so hard to tackle. Section 3.2 describes the primary characteristics of the agents as well as the system in which they are placed. Section 3.3 describes the state transitions of agents.

### 3.1 Black-Box and Glass-Box Views

In terms of modeling the occurrence of COVID-19 or any similar disease in a population, we find it useful to bring in the distinction between black-box and glass-box views of the epidemic.

The black-box view of the epidemic corresponds to what is observed about the incidence and spread of the disease in an affected population without widespread testing and tracing—agents just become ill with disease symptoms, and some need to be hospitalized, while others apparently stay healthy. Except in case of symptomatic cases, it is not known who all may carry the infection. This is related to, but not the same as, the standard definition of *black box epidemiology*: “the use of observational epidemiological methods and inference to arrive at conclusions about cause-effect relations between risk factors and disease” [28].

The glass-box view of the epidemic is the idealized condition where there is perfect knowledge of the extent of the epidemic, both in terms of what agents are infected, and the exact state that each agent is in. This too is not the same as the standard *white box epidemiology*, such as for example seen in the putative correlation between night-shift work and an increased risk of cancer [13].

As may be expected, in general far fewer agents are known to be infected in the black-box view, as asymptomatic infections are unseen. This leads to the situation depicted in Figure 2, where Figure 2a shows the black-box view, the limited perspective with relatively few agents showing symptoms, and Figure 2b shows the glass-box view, the real situation with many infected and exposed agents in the population.

**Figure 2:**
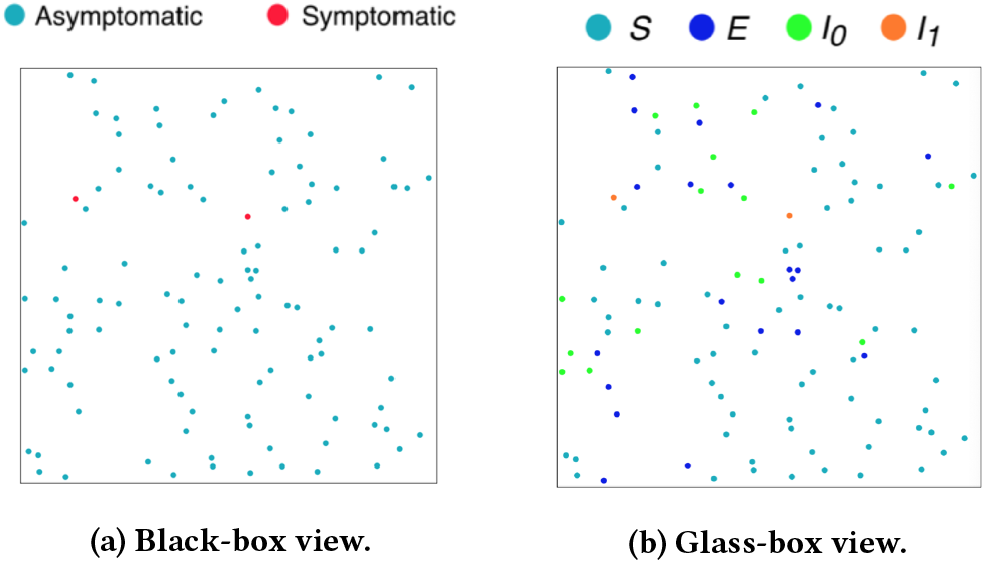
Two views of an epidemic at an early stage.

Another aspect of the difference between the black-box and glass-box views may be seen from Figure 3 which shows the apparent progress of the epidemic over time. The higher line corresponds to the glass-box view, where a much larger fraction of the population is seen to get the infection. The lower line corresponds to the black-box view, where a much smaller fraction of the population develops symptoms and is known to be infected.

**Figure 3:**
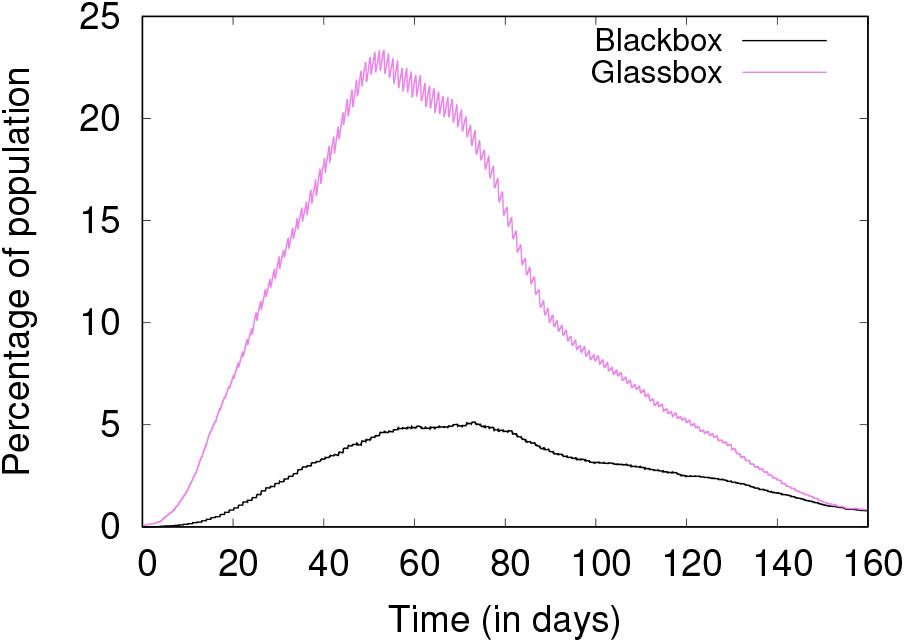
The Progress of an Epidemic, Glassbox vs. Blackbox Views.

The distinction between the black-box and glass-box views is made possible by the extended SEIR model depicted in Figure 1, which shows that the states *S, E, I*_0_, and *R* are all asymptomatic, and thus agents in these states appear not to be infected. An agent that is in state *I*_0_ is counted as infected in the glass-box view, but not in the black-box view. Agents that undergo the transition *S* → *E* → *I*_0_ → *R*, i.e., are exposed, infected, and subsequently recover without showing symptoms, and are not known to have been infected at all.

### 3.2 Individual and Social Attributes

We model humans in a society as a collection of agents, where each agent is a computational structure that captures some essential aspects of humans that are relevant to disease progression and spread. Some attributes are at an agent level, such as age, health, *etc*., while others are at a social level, such as the lockdown efficacy, social distancing, *etc*. We discuss the agent characteristics in Section 3.2.1 and the system parameters in Section 3.2.2.

#### 3.2.1 Agent Characteristics

An agent ϒ is formally defined as a five-tuple as follows:

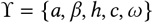

The agent attributes are:

- *a* is the age, 0 < *a* < 90.
- *β* denotes the level of hygiene, 0 ≤ *β* ≤ 1.
- *h* denotes the level of overall health, *h* ∈ {0, 1, 2, 3}.
- *c denotes* comorbidity level, *c* ∈ {−2, −1, 0}.
- *ω* denotes the immunity, *ω* ∈ {0, 1}.

These five attributes are further used to calculate the recovery score (*r*) and age score (*f*(*a*)), which are described in the following paragraphs, and are given by (1) and (2).

Age has been shown to be highly relevant to disease progression, with the elderly most at risk for worse outcomes [11].

The attribute *β* is included to model hygiene as seen in the real world in the form of protective masks, sanitizers, PPE’s, etc. This is particularly relevant in inter-personal transmission of the virus, which is described in detail in Section 3.3.1. A value of 1 corresponds to perfect hygiene and 0 corresponds to a complete lack of hygiene, with higher levels of hygiene correlated with lower chances of transmission.

We model health (*h*) as a 4-step discrete parameter from 0 to 3. A value of 3 denotes an agent in perfect health, and a value of 0 denotes an agent in poor health.

It is known that people with comorbid conditions have a higher death rate, and the death rate further varies based on the comorbid condition itself, with some conditions being worse than others [41]. A comorbidity (*c*) value of 0 implies the agent has no comorbid conditions, and a value of −2 implies the agent has the worst comorbid condition.

Immunity plays a major role in determining the speed of disease progression [19]. Weak immune response enables the virus multiply quickly and quickens the onset of symptoms and infectiousness. In our model, immunity can take a value of either 0 or 1, with 0 corresponding to an immunosupressed agent, and 1 corresponding to an agent with strong immune response.

We introduce recovery score (*r*) as an integrated metric that aggregates all these agent attributes, and directly correlates to the chances of recovery of an infected agent. Unlike all the other attributes, age has a very long range and therefore becomes difficult to combine with other attributes. Hence, age has to be transformed to a similar small range as the other attributes.

The effect of age on susceptibility to infection is quite complex [5, 15]. However, in the case of COVID-19, it is known that younger segments of the population (those below middle age) are generally less susceptible to infection, and that such susceptibility increases in middle age and becomes quite large for the elderly [11]. Based on this, we come up with a function (*f*) mapping the age of an agent with the infection risk, as follows:

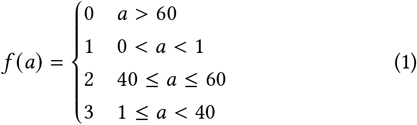

The age score function *f a* maps age, *a*, to a range of 0 to 3 where lower values correspond to higher risks. Using this age score in place of the actual age, we are able to get all the attributes in a similar range and direction, *i*.*e*., higher values of all attributes are a good thing, and lower values correspond to a higher risk of worse outcomes.

The values of these agent attributes are now added to get an integrated recovery score (*r*), which is a measure of the agent’s chances of recovery, and is given by:

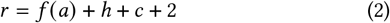

It is offset by 2 to counter the negative values of comorbidity and ensure the metric remains in the positive range. Hence, it can be seen that 0 ≤ *r* ≤ 8.

#### 3.2.2 System Parameters

In our model, we have five system-level parameters as given below:

- *α* denotes the efficacy of social distancing, 0 ≤ *α* ≤ 1.
- *κ denotes* the lockdown efficacy, 0 ≤ *κ* ≤ 1.
- *L* denotes the *lockdown* duration, in days. *L >* 0.
- *b* denotes the number of *beds* per 1000 agents, *b >* 0.
- Π denotes the fractional immunity in the system, 0 ≤ Π ≤ 1.

Social distancing has been widely used as a policy to slow the spread of disease in many places. There are many interactions between agents in a simulation, and social distancing is said to be followed in one such interaction if the agents are outside the infection ranges of each other. If social distancing is followed in a particular interaction, then there is no disease transmission due to the said interaction. We define the social distancing efficacy (*α*) as the ratio of the number of interactions where social distancing was followed to the total number of interactions.

Lockdown has been used as a strategy to various degrees by various countries to “break the chain” and slow the spread of infection. In a complete lockdown, all agents are static and their movements are completely restricted. However, more generally, not all agents observe lockdown and some continue to move around, and this is modeled through a lockdown efficacy parameter (*κ*). Another parameter of a lockdown is the duration, denoted by *L*. For example, a lockdown with *L* = 30 and *κ* = 0.8 implies there is a 30 day lockdown with 20% of agents still mobile, but the other 80% of agents are completely static.

To consider cases where a fraction of the population is immune to the disease, either from previous exposure and recovery, or by immunization or natural immunity, we define a parameter called *fractional immunity*, denoted Π, which gives the fraction of the agent population that is immune at the outset of a simulation. This notion of fractional immunity is similar to that seen in studies that seek to model the effect of a pre-existing or acquired immunity already present in a significant fraction of the population [17, 37]. Hospitals are considered to have two types of resources—normal beds and ICU beds. The normal beds are for agents in *I*_2_, and the ICU beds are for those in the more critical *I*_3_. The bed capacity of the system, denoted by *b*, is the number of beds per 1000 agents in the system.

### 3.3 State Transitions

The state transitions are as defined in Section 2.2 and their transition probabilities are described here. The probability of state transition from *S* to *E* is computed once for every interaction with an infectious agent as described in Section 3.3.1. However, the rest of the transition probabilities, given by (4)–(13), are all computed on a daily basis, and if none of the possible transitions occur on any given day, then the agent continues in the same state.

#### 3.3.1 Transition from S, the Susceptible State

Conventional methods of estimating disease transmission probability use differential equations and such tools, and work with populations rather than considering individual interactions [27, 45]. However, in an agent-based model, the probability of transmission cannot be constant and uniform across all interactions, and should be based on the interacting agents’ attributes.

A susceptible agent *A* in state *S* can get exposed to the virus through an interaction with another infectious agent *B*. If social distancing is followed in this interaction, then agent *A* is not exposed, and continues to remain in *S* state even after the interaction. However, if social distancing is not practiced, then the chances of transmission depend on the levels of hygiene (*β*) of both agents. The levels of hygiene of *A* and *B* may be denoted by *β*_*A*_ and *β*_*B*_ respectively. The probability of infection, denoted by *p* (*A, B*) is given by:

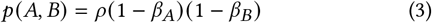

where 0 *< ρ* ≤ 1 is a scaling constant. If either agent practices perfect hygiene, transmission would not occur, and this holds in our model as well by (3). Otherwise, the agent *A* now transitions from *S* to *E* with a probability of *p* (*A, B*) due to the said interaction.

#### 3.3.2 Transitions from E, the Exposed State, and I_0_, the Asymptomatic State

The duration between an agent first reaching states *E* and *I*_1_ is termed as the *incubation period* (*λ*). The *latent period* (Φ) refers to the duration between states *E* and *I*_0_. In the case of COVID-19, the latent period is known to be smaller than or equal to the incubation period [30]. Accordingly, we fix the latent period is fixed such that 1 ≤ Φ ≤ *λ*.

The incubation period depends on a number of factors including host immunity (*ω*) and age (*a*) [19, 48]. The minimum, maximum and average values are denoted by *λ*_*min*_, *λ*_*max*_, and 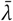 respectively. Current studies on COVID-19 indicate that the incubation period is between 2 and 14 days [9] with a mean of 5 days [29].

To calculate the transition probability for a weak immune agent with *ω* = 0, we use the following expressions. We sample a number *x* from a uniform distribution between 0 and 1. The agent transitions to *I*_1_ directly from *E* if the below condition is met:

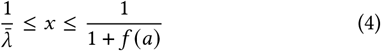

This creates a higher probability of transition to the *I*_1_ state if the age score *f* (*a*) is lower, since the range in which *x* is allowed to fall now has larger window based on the age score.

If an agent has strong immunity (*ω* = 1), it has a lower chance of directly transitioning into the state *I*_1_, as given by the following:

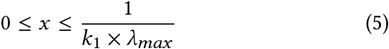

If the agent is in the *E* state, there is a chance of moving to state *I*_0_. For both weak and high immune agents, we use the following expression. We sample another number *y* from the same uniform distribution, and calculate the transition for *I*_0_ by:

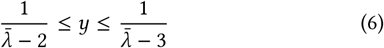

Sakurai *et al*. [40] indicate that the duration to resolution of infection for asymptomatic individuals is in the range of 3 to 21, and indicate that a considerable portion of asymptomatic individuals recover within 15 days. In our model, if an agent in *I*_0_ does not transition to *I*_1_ within 12 days, then it is considered recovered and moves to *R*.

#### 3.3.3 Transitions from I_1_, the Symptomatic State

A symptomatic infectious agent in *I*_1_ can transition to *I*_2_, *D*, or *R*, as seen in Figure 1. Multiple factors such as the recovery score (*r*) given by (2), the number of days spent in 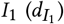, and the availability of hospital care, determine the next transition.

If hospital beds are available, the agent transitions from *I*_1_ to *I*_2_ with a probability given by:

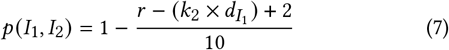

The probability of transition from *I*_1_ to *I*_2_ depends on the recovery score (*r*) and time spent in state 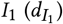. If *r* is high, it is less likely for the agent to move to state *I*_2_. If an agent spends more time in *I*_1_, then the probability of transitioning to *I*_2_ is greater. So, we use both parameters to calculate the probability *p*(*I*_1_, *I*_2_). We use a multiplicative constant *k*_2_ to rescale 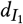 to be in the same range as *r*. As this is a probability, and hence should be between 0 and 1, the expression is scaled and offset appropriately.

However, if a hospital bed is not available, then the agent probabilistically transitions to death, *D*, as given by:

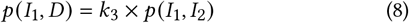

Also, if the agent remains in *I*_1_ for more than 12 days, then it recovers and transitions to *R*. This is with a small margin over the 10 days of infectivity indicated by the CDC [8].

#### 3.3.4 Transitions from I_2_, the Hospitalized State

An agent in *I*_2_ can transition to *I*_3_, *I*_1_, or *D*. These transitions also depend on the recovery score (*r*), duration in 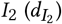, and the availability of ICU if needed. If ICU care is available, then the agent transitions from *I*_2_ to *I*_3_ with a daily probability given by:

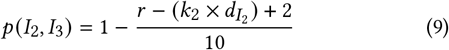

This equation is very similar to (7), and the scaling is done similarly as well.

If ICU care is not available, then the agent transitions to *D* with a probability given by:

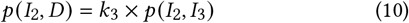

The daily probability that the agent transitions to *I*_1_ and becomes better is given by:

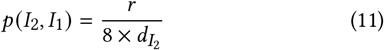

“Prolonged Length-of-stay (PLOS) [in hospital] is associated with increased mortality and other poor outcomes” [32]. In line with this, the probability of an agent in *I*_2_ going back to *I*_1_ should be inversely proportional to the hospitalized duration 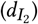, as seen in (11).

#### 3.3.5 Transitions from I_3_, the Critical State

An agent in *I*_3_ can either transition to *D* or *I*_2_, and this depends on the recovery score (*r*) and the duration in ICU 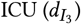.

The agent transitions to death with an increased probability as the time spent in ICU increases; a lower value of recovery score also implies a higher risk of death. Therefore, the daily probability of dying is given by:

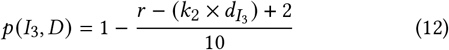

This is similar to the worse transitions from *I*_1_ and *I*_2_ and the same scaling factors are used here as well.

The daily probability of progressing and moving back to *I*_2_ is given by:

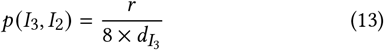

Agents with higher recovery score have better chances of recovering, and hence *p* (*I*_3_, *I*_2_) is proportional to *r* as seen in (13).

## 4 EXPERIMENTS AND RESULTS

In this section we discuss the experiments we have performed with agents and the disease model we have described previously.

### 4.1 Simulation Environment

Our simulation experiments are performed by circular agents in a 2-D enclosed planar environment. The environment scales based on the number of agents. We have performed all our simulation with 20,000 agents. Our simulations run for 160 simulated days, where 500 simulation cycles corresponds to 1 simulated day, and thus makes for a total 80,000 cycles for one experiment.

The initial range of values of each agent attribute is presented in Table 1. It is of course not possible to claim that these (or any other) simulation settings precisely describe all real scenarios, but wherever possible, we have calibrated the model to be close enough to real parameters as known from prior sources [11, 19, 41, 50].

**Table 1:** Notation. The presence of a dash indicates that there is no pre-set range for that parameter.

| Variable | Symbol | Range |
| --- | --- | --- |
| Hygiene | $\beta$ | [0,1] |
| Hygiene popn. mean | $\bar{\beta}$ | [0,1] |
| Age | $a$ | [0,90] |
| Health | $h$ | {0,1,2,3} |
| Comorbidity | $c$ | {-2,-1,0} |
| Immunity | $\omega$ | {0,1} |
| Social Distancing | $\alpha$ | [0,1] |
| Fractional Immunity | $\Pi$ | [0,1] |
| Lockdown efficacy | $\kappa$ | [0,1] |
| Transmission constant | $\rho$ | 0.25 |
| Incubation period | $\lambda$ | [1,14] |
| Latent period | $\Phi$ | [1,14] |
| Recovery score | $r$ | {0, 1, 2, ..., 8} |
| Bed capacity | $b$ | $0 \leq b \leq 1000$ |
| Duration in $I_*$ | $d_{I_*}$ | — |
| Scaling constants | $k_*$ | — |

For every experiment a population of agents is initiated and unique properties are assigned based on the range and distributions described in Table 1. Most agents are in the susceptible state (*S*) initially, but a small fraction of agents are in *I*_0_ and begin propagating the disease (see Section 4.2).

Each agent has attributes as described in Section 3.2.1 and the age score and recovery score are calculated using (1) and (2) respectively.

In each simulation cycle, every agent is triggered and follows Brownian motion (as used to model human crowd movement [23, 39]) and updates its location in the environment. Each agent has an identical infection range around it. Because of random motion, if any susceptible agent comes within this range of an infectious agent, there is a chance that infection can be transferred to the susceptible agent. This chance depends upon the level of personal hygiene *β* for both agents, as calculated by equation (3). Similarly at every trigger, each agent’s state transition probabilities are calculated and updated as described in Section 3.3.

Given some policies applied in a given day, some of the agent behaviors are controlled by said policies. For example, during a lockdown, the location of the agents is not updated; when social distancing is observed, an agent’s location is not permitted to be within the infection range of other agents.

To simulate isolation during quarantine we remove agents from the environment and keep them in a different quarantine environment for 14 days. Once they are recovered or upon completion of this duration, they are again sent back to the main environment. Deceased agents are removed from the environment completely. All our simulations not only produce output data, but also can be visualized as seen in the Figures 2a, 2b.

We use the MASON [31] simulation library, a Java simulation toolkit that scales well with a large number of agents and also helps in generating visuals of environment and agents. We used AWS C3 (compute optimised) Linux VMs with 4-core CPU and 8GB RAM.

### 4.2 The Base Case

This corresponds to the unrestricted spread of a contagion through an enclosed, initially completely-susceptible population. Even in this case, symptomatic infected (*I*_2_, *I*_3_) agents are isolated from the rest of the population. The number of isolated agents depends on hospital bed and ICU bed availability. Otherwise everything goes on as usual, with no restrictions on movement, no social distancing and no lockdowns.

One way to instantiate the base case is to admit a fixed number of infected agents into the population. The agents’ ages are sampled from a triangular distribution with a peak of 25, and a range of 0 to90. The mean hygiene level of the population is 0.6. At random, 3% of the population is assigned weak immunity in line with a study conducted in the US [18]. To start the onset of infection, we begin with .005% of infected agents already in the system. The incubation period is kept to a maximum of 14 days.

In the baseline as well as in other cases, the default hospital bed availability is 2 beds/1000 population [50]. We assume the availability of 1 ICU bed per 20 hospital beds.

Figure 4 shows the percentage of agents in each infectious state versus time, for the base case. We can see that the peak of asymptomatic (*I*_0_) cases is reached just before day 60. Similar peaks are seen for *I*_1_, *I*_2_, and *I*_3_ cases but with a delay, as observed in reality. The load on the public health infrastructure and the peak demand of critical care can be understood from the *I*_2_ and *I*_3_ curves. On day 120, the demand for hospitalization peaks, and roughly 1% of the entire population requires hospitalization at this stage.

**Figure 4:**
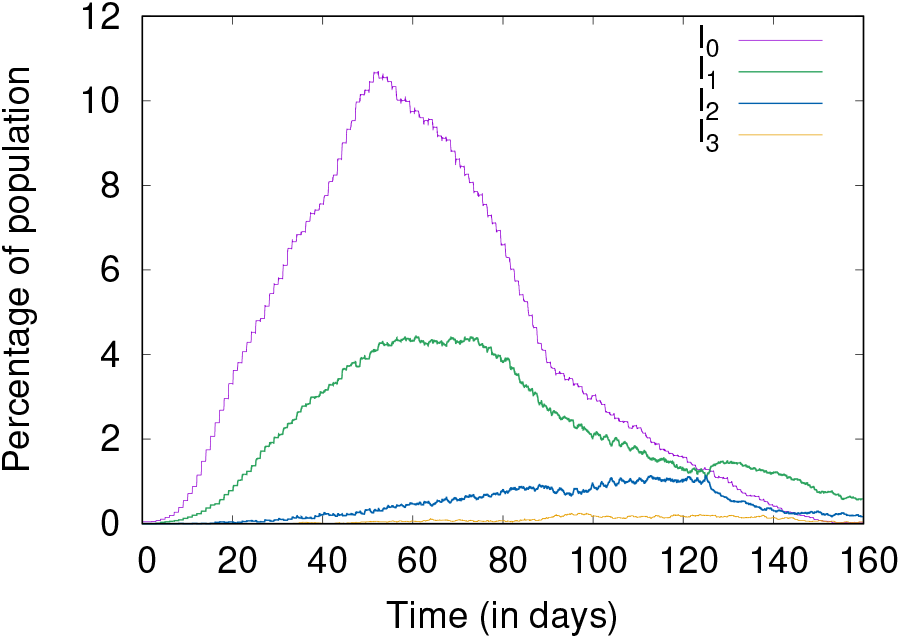
Baseline.

Figure 5 shows the cumulative numbers of infected, recovered, and deceased, also for the base case. Without any social distancing or other measures, the disease spreads fast, and nearly every agent in the population is eventually infected, and most subsequently recover by day 160. It can also be seen that only about 30% of the agents show symptoms even though the entire population was infected at one time or another. This shows that most agents (70%) progress to the *I*_0_ (asymptomatic) state but no further, as seen in reality as well [36].

**Figure 5:**
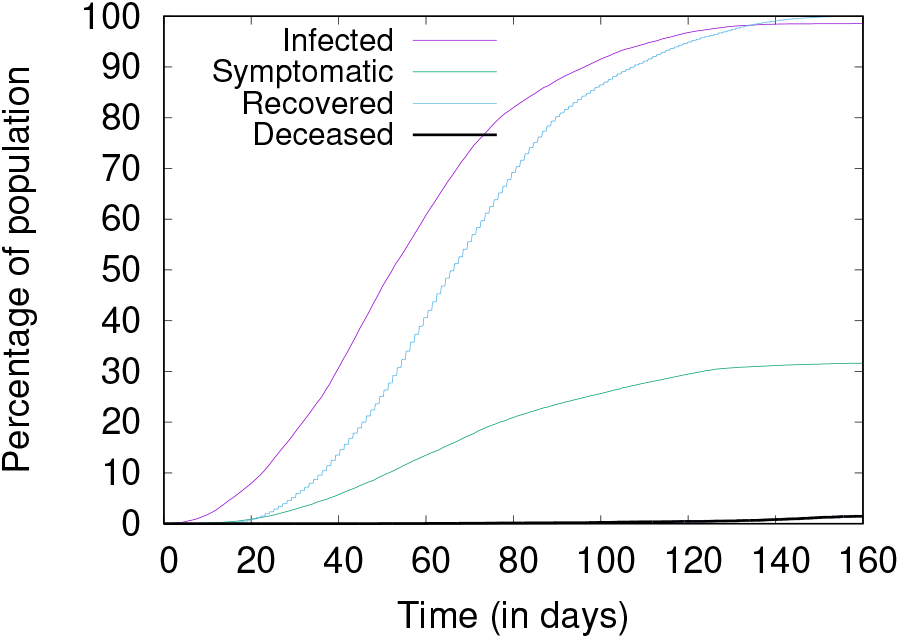
Cumulative figures of infection, recovery, and death.

### 4.3 Hygiene and Social Distancing

It has been repeatedly said that maintaining personal hygiene and social distancing are the key to slow down the spread of COVID-19. In this experiment, we compare the effects of two strategies—(i) a a high value of average personal hygiene with moderate social distancing 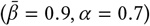; and (ii) a high average social distancing with moderate personal hygiene 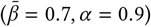.

Through simulation, we find that social distancing is more important than personal hygiene. As seen in Figure 6, the higher social distancing scenario reduces the peak load by about 50% when compared to the higher personal hygiene scenario. However, it is seen that both these cases are much better than the baseline.

**Figure 6:**
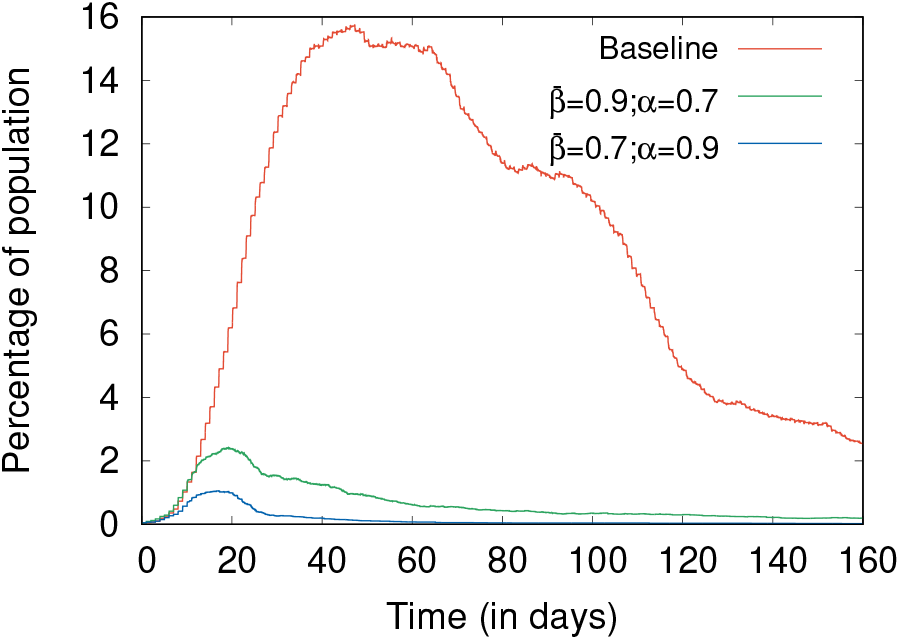
Comparing high hygiene and high social distancing.

Varying both mean personal hygiene and social distancing efficacy in the range of 0.4 to 0.9, we ran 10 simulations for each pair to understand the peak values and when they occurred. The average peak values and the median of days on which these peaks occur are shown in Table 2. With increasing mean hygiene 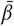, the peak days are delayed for all infectious states. It can be seen that increasing both 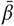 and *α* reduces the number of deaths, as expected. However, there is a bigger reduction in the number of deaths for every 0.1 increase in *α* as compared to the same 0.1 increase in 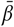. A similar trend is seen in the peak values also. The number of deaths is negligible for high values of *α* such as 0.8 and 0.9 even with lower values of 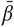, but similar high values of 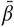 with lower *α* do not reduce the deaths to the same extent. Once again, this shows that social distancing is more important than personal hygiene.

**Table 2:** Peak values and deaths for different values of hygiene and social distancing.

| $\beta$ | $\alpha$ | Peak values (%) | | | Median peak day | | | $D$ (%) |
| --- | --- | --- | --- | --- | --- | --- | --- | --- |
| | | $I_1$ | $I_2$ | $I_3$ | $I_1$ | $I_2$ | $I_3$ | |
| 0.40 | 0.40 | 3.12 | 0.97 | 0.24 | 76 | 148 | 138 | 0.53 |
| 0.40 | 0.60 | 1.73 | 0.60 | 0.12 | 119 | 157 | 152 | 0.18 |
| 0.40 | 0.80 | 0.02 | 0.01 | 0.01 | 25 | 43 | 27 | 0.00 |
| 0.40 | 0.90 | 0.01 | 0.00 | 0.00 | 14 | – | – | 0.00 |
| 0.60 | 0.40 | 2.60 | 0.88 | 0.21 | 83 | 157 | 146 | 0.46 |
| 0.60 | 0.60 | 1.14 | 0.41 | 0.10 | 135 | 158 | 148 | 0.13 |
| 0.60 | 0.80 | 0.02 | 0.01 | 0.00 | 20 | 18 | – | 0.00 |
| 0.60 | 0.90 | 0.01 | 0.00 | 0.00 | 14 | – | – | 0.00 |
| 0.80 | 0.40 | 2.33 | 0.76 | 0.17 | 107 | 158 | 149 | 0.29 |
| 0.80 | 0.60 | 0.56 | 0.18 | 0.05 | 158 | 155 | 152 | 0.06 |
| 0.80 | 0.80 | 0.01 | 0.00 | 0.00 | 16 | 3 | – | 0.00 |
| 0.80 | 0.90 | 0.01 | 0.00 | 0.00 | 10 | 4 | – | 0.00 |
| 0.90 | 0.40 | 2.39 | 0.76 | 0.15 | 118 | 158 | 149 | 0.25 |
| 0.90 | 0.60 | 0.51 | 0.16 | 0.04 | 153 | 152 | 154 | 0.04 |
| 0.90 | 0.80 | 0.02 | 0.00 | 0.00 | 13 | 6 | – | 0.00 |
| 0.90 | 0.90 | 0.01 | 0.00 | 0.00 | 11 | 3 | – | 0.00 |

### 4.4 Lockdowns

Lockdown is another strategy that has been used in some form across many countries to combat the spread of COVID-19. In a set of experiments to study its effects, when lockdown is applied, agents are immobile. In practical settings, perfect lockdowns are impossible (due to some people being essential workers, or defiant of the lockdown policy), so we simulate partial lockdowns using a parameter of lockdown efficacy (*κ*), which is the fraction of agents who follow the guideline fully and stay completely immobile during this period. But even immobile agents can always come in contact with other agents who are mobile and not observing the lockdown policy. We experiment with different values of *κ* to see its effects on the infection spread.

We run experiments of imposing a 30-day lockdown, starting on day 10, with varying efficacy. In Figure 7 we can see that during a 30-day lockdown period the infection rises faster with *κ* = 0.7 compared to tighter and strict lockdown with efficacy 0.9. It also becomes apparent that once the lockdown is lifted, without any other policy such as increased hygiene level or social distancing, we get a worse peak than the baseline. This implies that a lockdown is not a complete solution, and can possibly only provide time to prepare for a later peak of infections. We can clearly see lockdowns are effective at delaying the peak. Even with *κ* = 0.7 which means 30% of agents do not follow any guideline, we do not see a high peak during the lockdown period.

If a partial lockdown is observed with (and followed by) higher social distancing and hygiene, there is some improvement. In Figure 8, we see that the proportion of infected agents stays at a moderately high level, but there is not a much higher peak unlike Figure 7.

**Figure 7:**
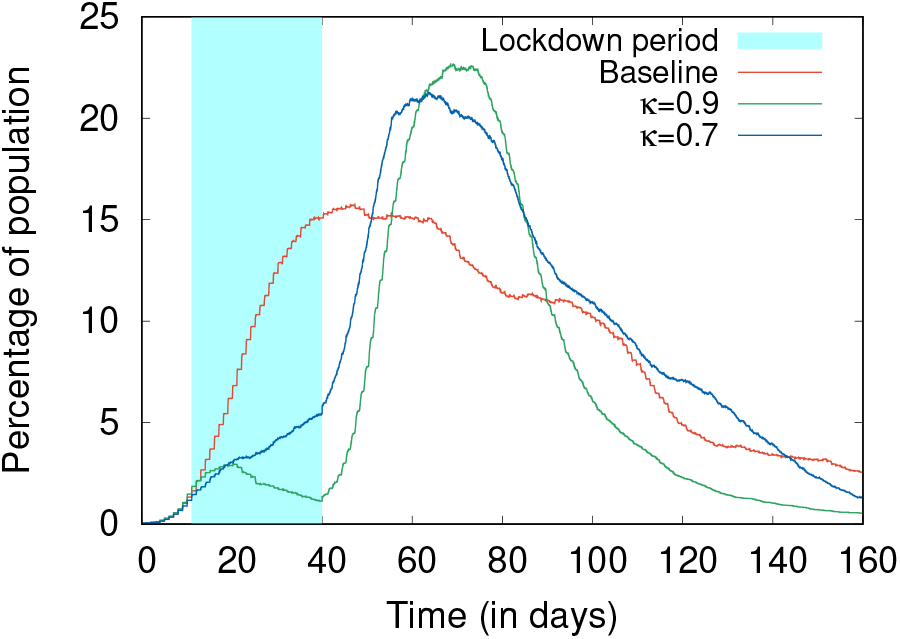
Effects of varying lockdown efficacy.

**Figure 8:**
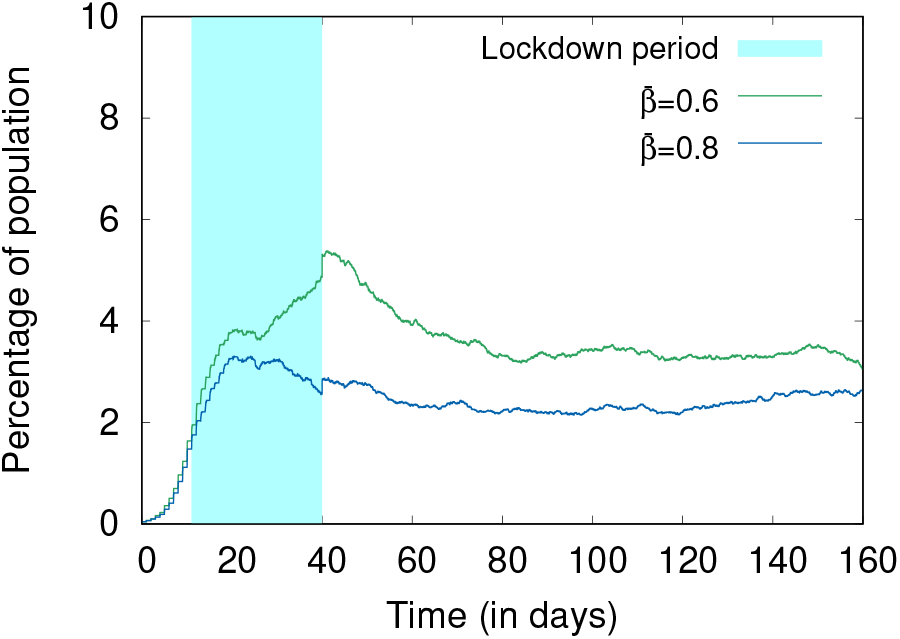
Effects of different levels of hygiene during and after lockdown with *κ* = 0.8; *α* = 0.7.

We also present the the effect of social distancing post lockdown. In Figure 9 we show the effects of varying levels of social distancing. Social distancing is a much more sensitive parameter compared to hygiene, so a small change in social distancing changes the number of infections significantly. We can see that with a higher value of *α* (0.8), the proportion of infected agents is much lower than with a moderate value (0.6).

**Figure 9:**
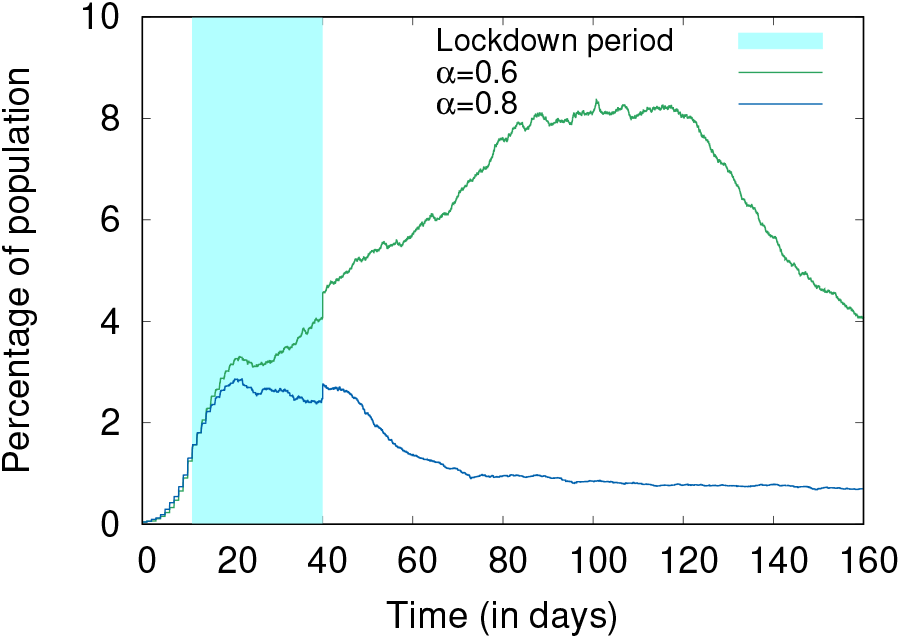
Effects of different levels of social distancing during and after lockdown with 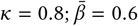.

### 4.5 Fractional Immunity

This set of experiments studies an initial state with some fraction of the population already immune to the virus (which can be achieved by natural immunity, post infection recovery, or by vaccination). We describe this as fractional immunity in certain population.

In one set of experiments we start the simulation with baseline conditions (no hygiene or social distancing), but varying Π, the fractional immunity of the agent population, from 0.2 to 0.8 (corresponding to 20% through 80% of the population being immune). Figure 10 shows that even when 20% and 40% of population is immune to the virus, we can have significant peaks in the infection in the absence of hygiene and social distancing. Even at 0.6 fractional immunity, there are a significant number of infections. It is only when there are 80% immune agents that we really see an effective control on the spread of infection.

**Figure 10:**
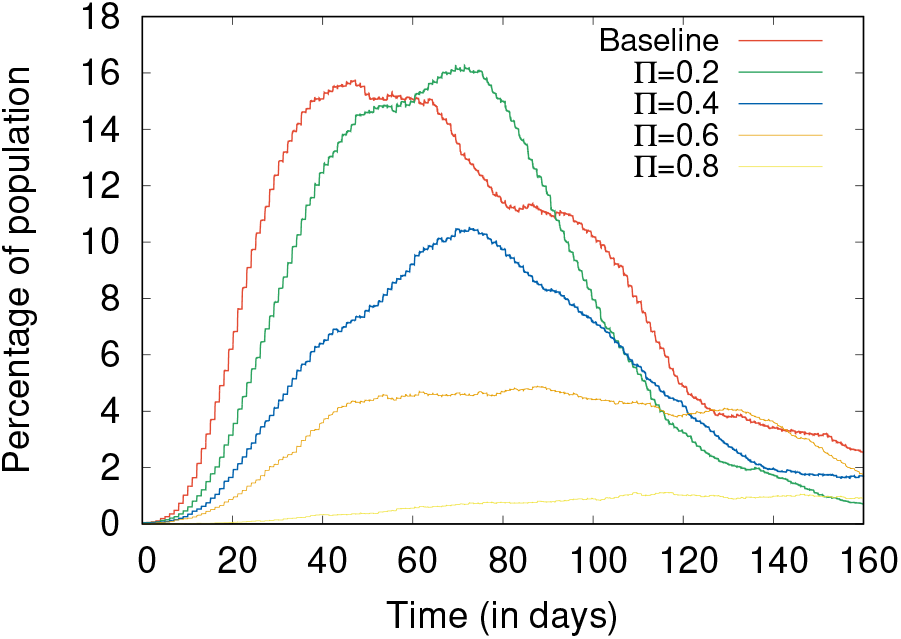
Effects of varying the levels of fractional immunity.

We experimented with varying levels hygiene and social distancing with 20% and 40% of population being immune.

Figure 11 shows the effect of varying social distancing, keeping 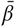 constant at 0.6, for Π at 0.2 and 0.4. As expected, the proportion of infected agents is lower with a higher value of Π (Figure 11a vs. Figure 11b). With a fractional immunity of 0.4 and moderate hygiene and social distancing, the value of the peak reached here is a fraction, roughly one-fourth, of the peak value seen for the same Π in Figure 10.

**Figure 11:**
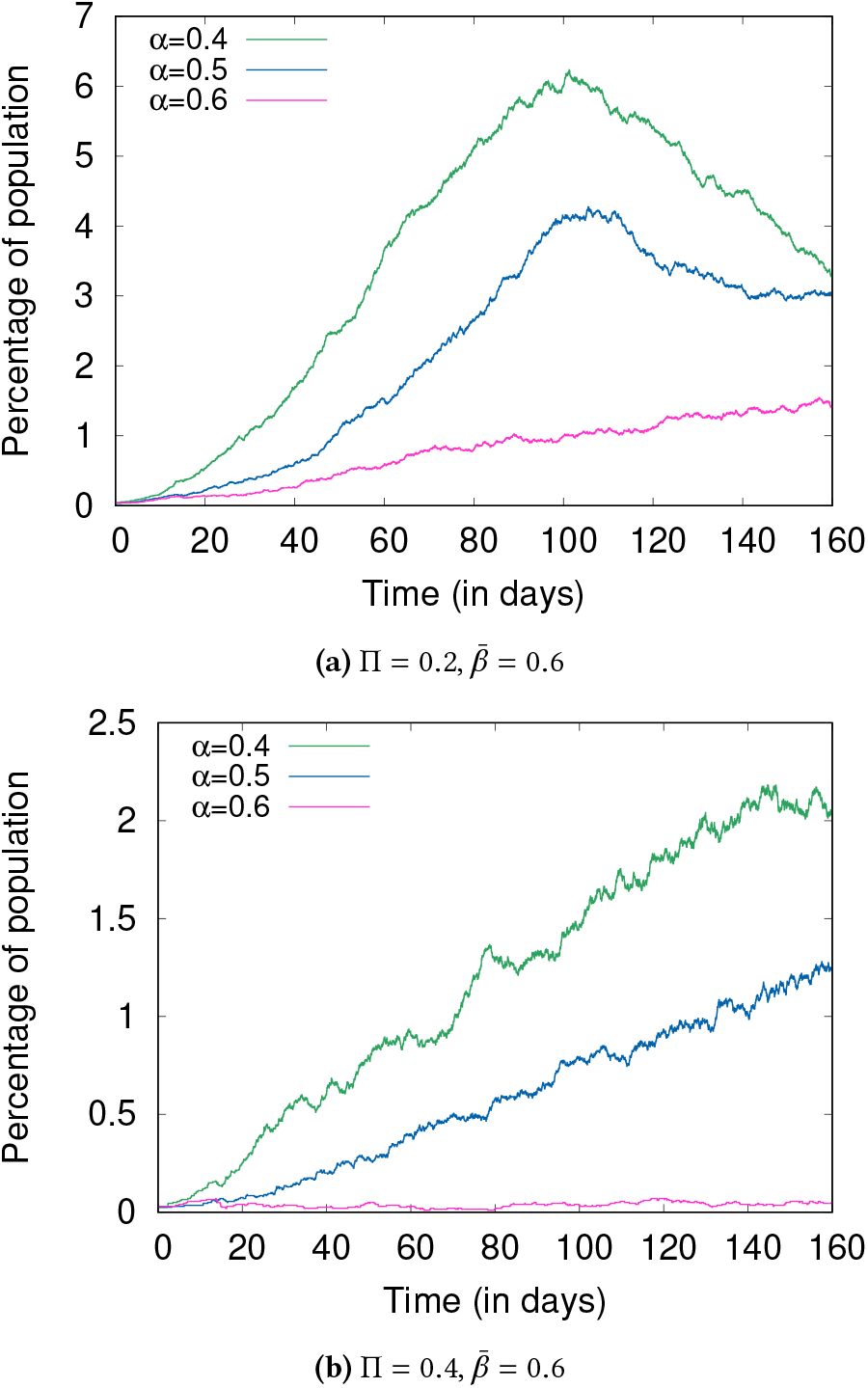
Comparing the effects of *α* at two different levels of Π.

**Figure 12:**
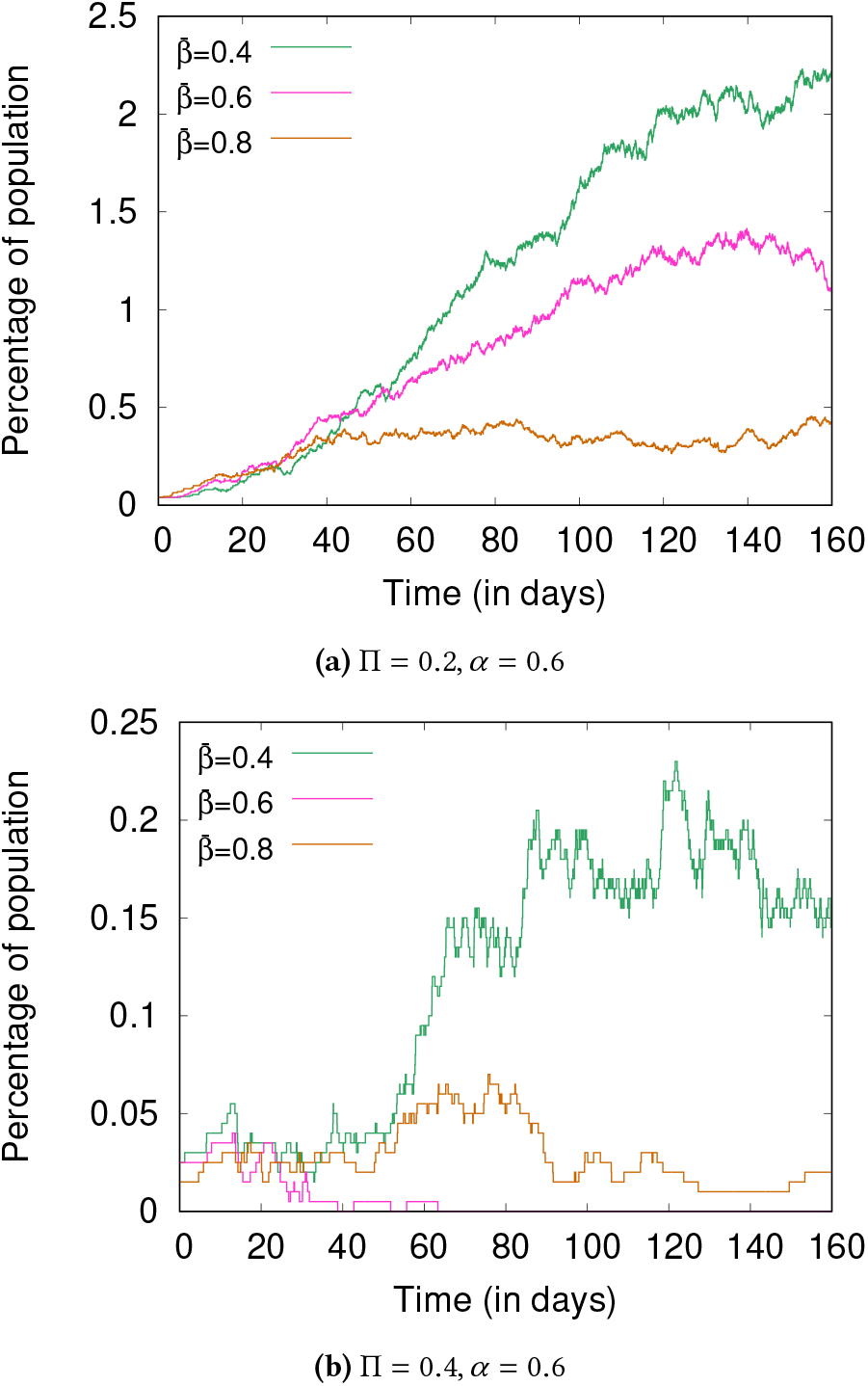
Comparing the effects of *β* at two different levels of Π.

Similarly, Figure 12 shows the effects of varying hygiene with the social distancing efficacy kept constant at 0.6. We see a similar trend with the number of infections decreasing with increasing hygiene.

### 4.6 Surges in Hospital Beds and ICUs

In this experiment, different levels of surge capacity are considered (50% increase, 100% increase in the bed capacity). On the day-50, these surge capacities are added to existing capacity. This is meant to reduce the rate of death on account of inadequate hospital capacity.

Figure 13 shows the percentage of deceased agents. We can see that a surge does reduce the number of deceased agents, but not in a linear fashion. The number of deceased agents drop by 10% by adding 50% more hospital and ICU beds, and further reduces by 20% when the number of beds and ICU units is doubled.

**Figure 13:**
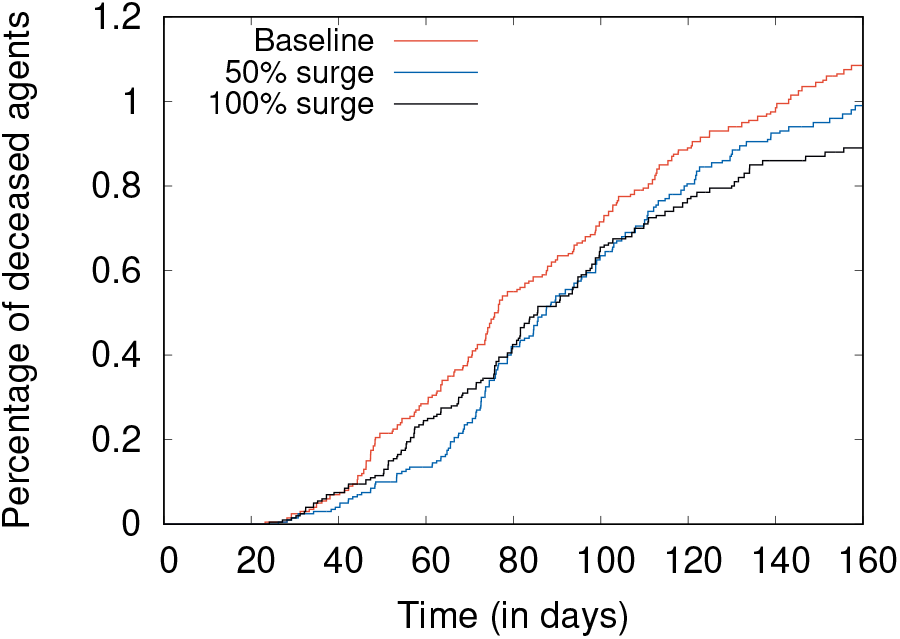
Common parameters across all three experiments: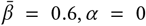. Baseline (*b* = 2), 50% surge (*b* = 3), 100 % surge (*b* = 4)

## 5 CONCLUSION

Data-driven approaches and purely statistical models for COVID-19 exist, but have some significant limitations [43]. This is in large part due to the mostly unfamiliar nature of the novel coronavirus SARS-CoV-2, and the fact that traditional equation-based models fail to capture the dynamics of systems that are heavily dependent on factors such as population distributions and agent characteristics. Such top-down models are also ill-equipped to show the emergent consequences that arise from fine changes in individual behaviors, and policy choices that cannot be readily expressed in mathematical form [20].

We therefore use an agent-based model for a qualitative assessment of the COVID-19 pandemic in an enclosed population where agents represent humans. This model is thus a bottom-up approach to capture the unpredictable interactions between members of a society, and understand the manner in which COVID-19 may spread under different conditions.

As COVID-19 is a new disease and its properties are still being discovered, it is possible that some assumptions of our model, which are in line with existing knowledge about the disease, may have to change. Even so, we believe that this work demonstrates some important emergent properties and helps analyze what-if scenarios.

Even with a moderate level of personal hygiene and social distancing being widely observed, the epidemic can be effectively contained even with a relatively small fraction (such as 20–40%) of the population being immunized by vaccination or prior exposure and recovery. This means that dire prognostications that COVID-19 will eventually infect a majority of the population are fortunately unlikely.

Prior agent-based models for COVID-19, such as by Silva *et al*. [44] and Cuevas [10], deal in a narrower set of possible circumstances. Our work is the first to give conclusions about lockdowns and behavioral choices such as personal hygiene and social distancing, and indicate the likely progress of the disease under specific conditions. It is our hope and expectation that this work will serve to illuminate certain aspects of the spread of the disease and show the value of certain types of control measures that may be applied. It should thus be of use to policy makers and authorities as well as the public at large.

## Data Availability

The code and documentation for this work can be accessed from https://github.com/ABM-for-Covid/ABM-for-Covid-19. We have also created an interactive application (https://abmforcovid.org) for anyone to run experiments and test with their own strategies.

## Notes

### Competing Interest Statement

The authors have declared no competing interest.

### Clinical Trial

Not applicable

### Funding Statement

No external funding was received for this work.

### Author Declarations

Not applicable as this work uses simulations but no clinical or animal experiments.

### Summary of Updates

Minor typographic and such corrections only.

